# Managing the risk of a COVID-19 outbreak from border arrivals

**DOI:** 10.1101/2020.07.15.20154955

**Authors:** Nicholas Steyn, Michael J. Plank, Alex James, Rachelle N. Binny, Shaun C. Hendy, Audrey Lustig

## Abstract

In an attempt to maintain elimination of COVID-19 in New Zealand, all international arrivals are required to spend 14 days in government-managed quarantine and to return a negative test result before being released. We model the testing, isolation and transmission of COVID-19 within quarantine facilities to estimate the risk of community outbreaks being seeded at the border. We use a simple branching process model for COVID-19 transmission that includes a time-dependent probability of a false negative test result. We show that the combination of 14-day quarantine with two tests is highly effective in preventing an infectious case entering the community, provided there is no transmission within quarantine facilities. Shorter quarantine periods, or reliance on testing only with no quarantine, substantially increases the risk of an infectious case being released. We calculate the fraction of cases detected in the second week of their two week stay and show that this may be a useful indicator of the likelihood of transmission occurring within quarantine facilities. Frontline staff working at the border risk exposure to infected individuals and this has the potential to lead to a community outbreak. We use the model to test surveillance strategies and evaluate the likely size of the outbreak at the time it is first detected. We conclude with some recommendations for managing the risk of potential future outbreaks originating from the border.

## Introduction

The COVID-19 outbreak originated in Wuhan China in late 2019 before spreading globally to become a pandemic in March 2020 (1). Many countries experienced widespread community transmission after undetected introductions of the disease. This led to rapid growth of new infections in many countries. New Zealand experienced an outbreak of COVID-19 between March and May 2020, but successfully controlled the epidemic and ultimately eliminated community transmission of the virus for over 100 days (2, 3). During this time, the country’s border remained closed to everyone except citizens and residents, with a mandatory 14-day period of government-managed quarantine for all international arrivals. Countries such as Iceland, Taiwan, Vietnam and some states of Australia have reduced cases to very low numbers and are managing the virus with a combination of border controls and domestic restrictions. Other countries managing ongoing epidemics, including several European countries, also use border controls including quarantine to reduce the number of imported cases.

The goal of quarantining international arrivals differs depending on a country’s strategy for managing COVID-19. For countries with ongoing epidemics, such as the UK, intercepting 95% of imported cases and preventing them from infecting others in the community would be considered a good outcome (4). However, for countries with an elimination strategy, such as New Zealand, or a goal of maintaining very low prevalence of COVID-19, interception rates need to be as high as possible to minimise the risk of seeding new community outbreaks. This strategy also requires a robust re-emergence plan for dealing with new community cases and acting rapidly to contain the spread of the virus to avoid a major outbreak. This includes rapid testing, case isolation and contact tracing, as well as the option of local or regional restrictions should significant new clusters occur.

Government-managed quarantine facilities pose a risk of seeding community outbreaks. For example, the Australian state of Victoria experienced a major outbreak in July 2020 (5), thought to have originated from close contacts between people staying in government-managed quarantine and people working there. In New Zealand, there was a community outbreak in August 2020 that likely originated at a quarantine facility, and at least six instances between August and December 2020 of either a quarantine worker being infected with COVID-19 or a recent international arrival entering the community while still infectious. These events demonstrate the importance of minimising the risk to frontline border-facing workers and at all points of contact between international travellers and the community. The people working in these roles include securely-employed, highly-trained health professionals, who administer regular COVID-19 swab tests and health checks, full-time defence staff who manage security and logistics, through to less-secure contract staff who clean or provide transport to quarantine facilities. These facilities and the individuals staying and working in them are part of the front line of defence against a reintroduction of COVID-19. It is essential to understand and minimise the risks associated with them to prevent future community outbreaks.

Mathematical models of the spread and control of COVID-19 have been used successfully in informing the New Zealand government response to the pandemic (6-8). Wilson, Baker (9) modelled the effects of different policies, including compulsory mask use and scheduled RT-PCR tests for travellers, to mitigate the risk of quarantine-free travel between two countries with very low prevalence of COVID-19, such as New Zealand and Australia. Wilson, Schwehm (10) considered the levels of community surveillance at healthcare facilities that would be required to detect new outbreaks early enough to stop a large outbreak. Previous studies have modelled effects of non-pharmaceutical interventions on transmission dynamics in countries where community outbreaks have already established. For instance, Aleta, Martin-Corral (11) showed that strict population-wide social distancing combined with robust contact tracing, testing and home quarantine could successfully maintain transmission of SARS-CoV-2 at levels that do not overwhelm healthcare systems.

In designing border measures to prevent reintroduction of COVID-19, there are two main sources of risk that need to be considered. Firstly, there is a risk that a recent international arrival could enter the community while infectious. Secondly, there is a risk that a frontline border-facing worker, for example someone working in a quarantine facility, airport or port, could become infected and seed a community outbreak. These are low-risk, high-consequence events: they happen rarely but when they do happen they pose a major threat to countries, such as New Zealand, where restrictions on community activities and gatherings have been largely relaxed. Mathematical modelling is an effective tool in this situation because, in the absence of direct data on the risk of rare events, models can synthesise data on underlying aspects of the process (e.g. transmission dynamics, virus incubation period, surveillance programs, and false negative testing rates) to quantify this risk (12). We use a simple mathematical model of COVID-19 transmission and testing to investigate the effect of different border policy settings on the risk of a community outbreak seeded at the border via each of these routes. Our model takes account of realistic false negative rates for RT-PCR tests (13).

For the risk posed by international arrivals, we compare a testing-only policy with varying combinations of mandatory quarantine and testing schedules. We also propose a novel metric that can be used to estimate the level of transmission between individuals in managed quarantine facilities. For risk posed by frontline border workers, we investigate the effect that different testing schedules have on the likely size of an outbreak at the time of first detection. We consider two situations: one where the outbreak is first detected in the frontline worker, and one where it is first detected in a secondary case (i.e. a contact of a frontline worker) or later. This serves to quantify the risk of a serious outbreak depending on how the outbreak is first detected. We also estimate the risk that an infected individual has travelled from the region in which the outbreak was detected to another region before the outbreak was detected.

We conclude that 14-day quarantine with two scheduled tests for all international arrivals is a robust system for preventing imported cases entering the community. Steps should be taken to minimise opportunities for transmission between individuals in quarantine facilities, which can substantially elevate the risk. All frontline border-facing workers should be tested weekly to increase the likelihood that an outbreak seeded by a frontline worker is detected before it grows very large. Our analysis also provides a framework for supporting decisions on the nature and scale of the response needed to control a newly detected community outbreak.

## Methods

This section describes models used to simulate the COVID-19 spread and testing in different scenarios. First, we define the model for the transmission of COVID-19 and specify when individuals are tested for COVID-19, and their probability of returning a positive test result. We then describe the initial conditions and parameter values for these models to represent two different scenarios. Scenario 1 models transmission among international arrivals in a quarantine facility to estimate the probability of an individual entering the community while still infectious under different quarantine and testing regimes. Scenario 2 models a community outbreak seeded by a frontline quarantine worker. Finally, we define a simple model for calculating the probability that an infected individual has travelled outside a given region at the time an outbreak is detected.

### Transmission and testing model

#### Transmission model

We use a continuous-time branching process to model the spread of COVID-19 (6); see Table 1 for parameter values. We assume that 33% of infections are subclinical and that subclinical infections are 50% as infectious as clinical cases (14-16). Each infected individual *i*causes a randomly generated number *N*_*i*_∼*NegBin*(*R*_*i*_,*k*)of new infections, where *R*_*i*_=2.5 for clinical community cases and *k* is the overdispersion parameter (17). Times of secondary infections relative to infection of the index case are independent random variables drawn from a Weibull distribution, with mean and median equal to 5.0 days and standard deviation of 1.9 days (18). This distribution represents a transmission rate that is a continuous function of time since infection (Figure 1). The model does not explicitly include a latent period (i.e. period in which an individual is infected but not yet infectious). However, the shape of the Weibull generation time distribution means that there is a low probability of onward transmission occurring in the first few days after infection. The time taken to develop symptoms is gamma distributed with mean 5.5 days and standard deviation 2.3 days (19).

**Table 1.**
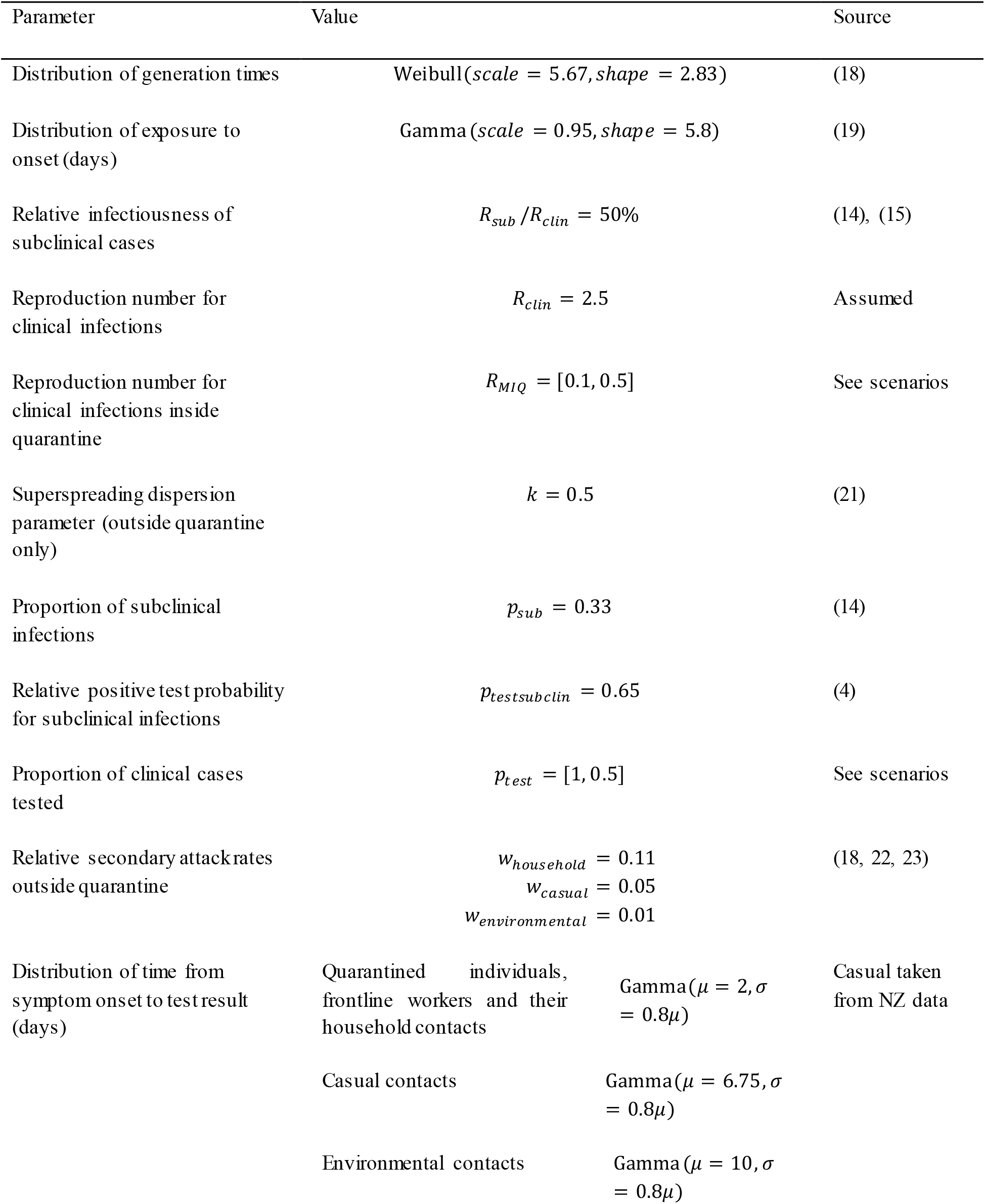
The parameters used in the model and their source.

**Figure 1.**
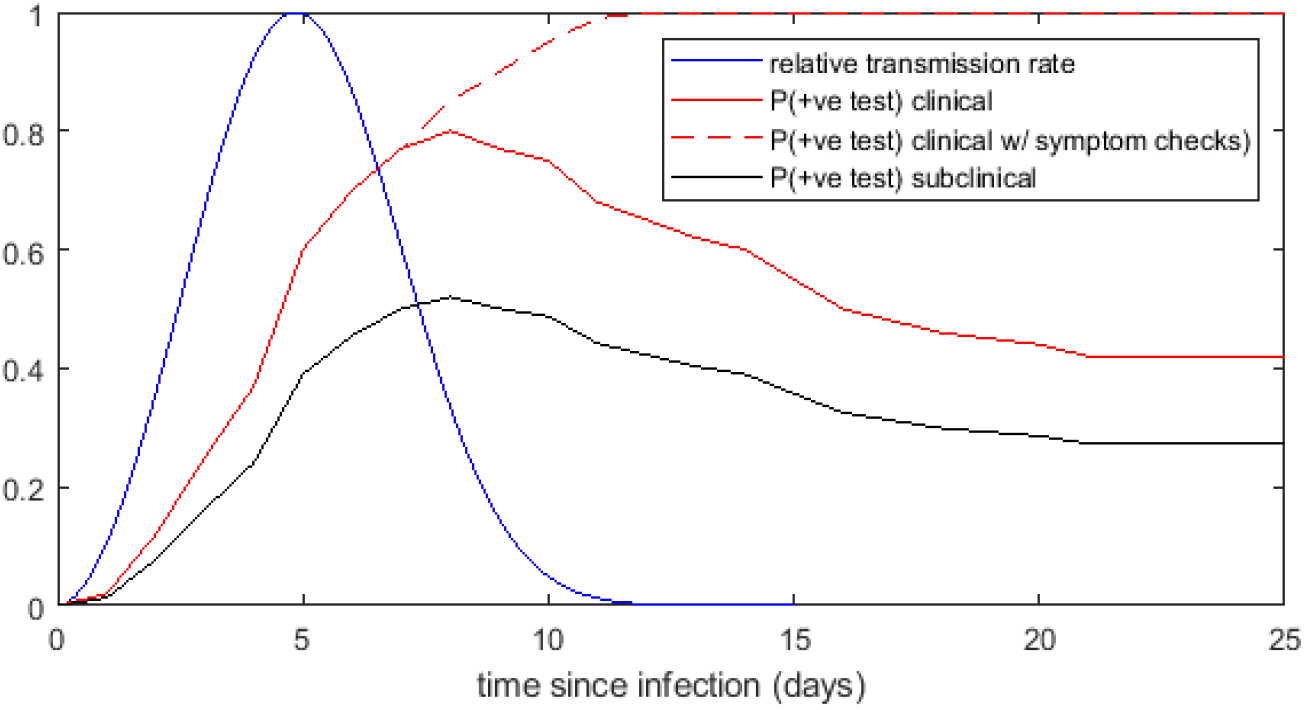
Model transmission rate relative to peak transmission rate (blue), probability of testing positive by RT-PCR test for clinical cases (solid red), probability of testing positive by RT-PCR test with additional symptom checks by a health professional (dashed red), and probability of testing positive by RT-PCR test for subclinical infections (black), as a function of time since infection. The transmission rate function is a Weibull distribution with mean 5.0 days and standard deviation 1.9 days (18). The probability of testing positive by RT-PCR test is based on the results of (13). The addition of symptom checks is assumed to increase the probability of testing positive due to diagnosis of probable cases based on clinical symptoms.

#### Testing model

Testing for COVID-19 occurs for one of two reasons in the model: (1) testing triggered by onset of symptoms; and (2) scheduled testing regardless of symptoms of either recent arrivals in quarantine or frontline workers in quarantine facilities. Assumptions about test sensitivity and the criteria and timing for symptom-triggered testing are described below. We investigate different regimes for scheduled testing of travellers and quarantine workers. There is no scheduled testing of general community members.

#### Test sensitivity

Both symptom-triggered and scheduled tests are assumed to be RT-PCR tests with a sensitivity (i.e. probability of correctly returning a positive result) that depends on the time since infection (13). The sensitivity is very low in the first 3 days after infection and is at its highest (approximately 80% for clinical cases) 8 days after infection (Figure 1). In some scenarios, we investigate the effect of supplementing tests with symptom checks carried out by a health professional. We assume that, when carried out more than 8 days after infection, symptom checks increase the probability of a case being identified relative to a RT-PCR test alone, with a 100% sensitivity more than 10 days after infection (Figure 1). This assumption reflects diagnosis of “probable” cases based on clinical assessment in the absence of a positive RT-PCR result. We assume that subclinical infections have a probability of testing positive that is *p*_*testsubclin*_= 65% of that of clinical cases (4), reflecting lower test sensitivity for mild infections. For simplicity, we assume that the results of multiple tests on the same individual are independent.

#### Symptom-triggered testing

Clinical cases receive a symptom-triggered test with probability *p*_*test*_ some time after onset of symptoms. Provided the test does not give a false negative, the case will be detected at that point. Subclinical infections remain asymptomatic or pauci-symptomatic throughout their infection and do not receive symptom-triggered tests. The mean time between symptom onset and a positive test result for New Zealand cases in the March to May 2020 outbreak was approximately 6 days (7). Managed isolation and quarantine facilities have measures in place for early identification of potential cases in frontline workers (e.g. regular health checks and additional training in infection, prevention and control requirements) and quarantined individuals (e.g. information on COVID-19 risk and regular health checks) (see Appendix for details). We therefore assume that individuals in quarantine, frontline workers and their families have an average of 2 days from symptom onset to test, whereas members of the general community have an average of 6 days from symptom onset to test (estimated from New Zealand data (7)). Secondary cases infected via environmental transmission are assumed to have very low awareness and have an average of 10 days from symptom onset to test. Here, environmental transmission means transmission via a contaminated surface or airborne particles (Vardoulakis, Sheel (20)) without direct contact, defined as exposure within 2 metres of a case during their infectious period without appropriate personal protective equipment. Processing of tests is assumed to be fast and results are received on the same day as testing. Due to variation between individuals, the high chance of a false negative test result, asymptomatic individuals and, in some scenarios, a low testing rate, the first detected community case may not be the seed case. The model does not consider community case isolation measures as we are interested only in the initial time period before the first case is detected and before contact tracing and isolation measures are initiated.

### Model scenarios

#### Scenario 1: international arrivals in a quarantine facility

We initialise simulations with a single seed case, representing an infected international arrival entering a quarantine facility. The infection time of seed cases is chosen uniformly at random in the 14 days prior to arrival. However, if the individual develops symptoms and subsequently tests positive prior to travel, they are assumed not to have travelled and are excluded from the sample. For scenarios with transmission between individuals in quarantine, we use a relatively small value for the within-quarantine reproduction number *R*_*MIQ*_(*R*_*MIQ*_= 0.1 for moderate transmission; *R*_*MIQ*_= 0.5for high transmission) to model the limited opportunities for contacts among individuals in a quarantine facility. We assume there is no superspreading within quarantine facilities, i.e. *N*_*i*_∼ *Poiss*(*R*_*MIQ*_) for a clinical case that spends their whole infectious period in a quarantine facility. Individuals that are infected during their stay in quarantine could be at any point in their stay, i.e. we assume there is no attempt to group returning travellers by arrival date.

In addition to symptom-triggered testing described above, all international arrivals undergo a scheduled test with symptom checks on day 3 and day 12 of their 14-day stay in quarantine. All cases that are detected in quarantine (whether via a positive test result or being diagnosed as a probable case) are moved to new facilities dedicated to housing confirmed and probable cases and with stricter infection prevention procedures (see Appendix for details). Modelling potential transmission within these separate facilities is beyond the scope of this study. We assume there is no onward transmission from cases once they have been detected, and they are completely isolated and not released until fully recovered. Cases that pass through quarantine undetected but are more than 14 days after infection are assumed to pose no further risk of onward transmission.

We simulate the model for alternative quarantine periods and testing regimes, and with different levels of within-quarantine transmission. The key model output is the proportion of simulations in which an individual is released into the community while still infectious (defined as being within 14 days of infection). The released infectious individual may be either an imported case or a case infected within the quarantine facility. Individuals released within 5 days of infection still have at least 50% of their onward transmission potential ahead of them (see Figure 1), and are defined as being highly infectious because they pose a greater risk of community transmission. Assuming multiple international arrivals are independent, our results can be interpreted as the risk per case arriving at the border.

We also record the proportion of all cases detected in quarantine that are detected during the second week after arrival. We investigate the relationship between the transmission rate in quarantine (measured by the within-quarantine reproduction number) and this proportion. Since the proportion of cases detected during the second week is readily observable, this may provide a practical way to infer whether significant transmission is occurring within quarantine facilities, which is difficult to observe directly.

#### Scenario 2: community outbreak seeded by a frontline worker

Simulations of community spread are seeded with a single infectious case representing a frontline border worker. We set the negative binomial overdispersion parameter *k* to be 0.5, which allows for heterogeneity in number of secondary cases and some superspreading (21). Infected individuals are randomly assigned an infection route depending on the type of contact: household (the index case and secondary case reside in the same household); work/casual (the index and secondary case have had direct contact outside the home, e.g. at work); or environmental (there has been no direct contact between the index and secondary case, and transmission is via a contaminated surface or airborne particles). These are weighted by the relative attack rates for each contact route, *w*_*route*_, based on the observation that within-household transmission accounts for 2-3 times more infections than outside-household transmission (22, 23), and the assumption of (18) that around 10% of all transmission is environmental. The model has no fixed household size as we are only interested in the very early stages before the outbreak is detected, where only one or two household members are typically infected. All secondary infections and their descendants are assumed to be members of the general community (i.e. we neglect the possibility of transmission from one frontline worker to another, or from a general community member to a frontline worker). Two community awareness scenarios are investigated: low community awareness where *p*_*test*_ = 50% of clinical infected individuals (including frontline workers) ever present for symptom-triggered testing; high community awareness, where *p*_*test*_ = 100% of clinical infected individuals are eventually tested. For each simulation, we calculate the total number of infected individuals *N*_*c*_ at the time the first case is detected.

### Inter-regional travel model

We use the results on expected outbreak size to estimate the probability that an infected individual has travelled outside a given region at the time the outbreak is first detected. The key model variables are the number of cases *N*_*c*_ infected before the first case was detected and the number of days *D*_*i*_(*i=* 1, …,*N*_*c*_) since each of these cases was infected. Together, these give the total number of opportunities for daily travel by any infected individual. The daily individual probability of travel is *r/M* where *M* is the population size of the region in which the outbreak was detected (region A) and *r* is the average number of individuals that travel to another specified region (region B) each day. The probability *P*_*travel*_ that an infected individual has travelled from region A to region B before the first case was detected is:

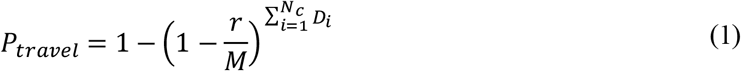

## Results

We run the model in the two scenarios described in Methods: (1) within a quarantine facility to estimate the probability an individual will leave the facility while still infectious; and (2) in the community external to quarantine facilities to estimate the size of an outbreak initiated by an infected frontline worker.

### Scenario 1: international arrivals in a quarantine facility

Various countries have mandated 14-day quarantine periods for some or all international arrivals (4). For example, all international arrivals to New Zealand are required to spend 14 days in a government-managed isolation and quarantine facility during which they are tested twice (around day 3 and day 12 after arrival) (24). No individual is allowed to leave quarantine without returning at least one negative test result. We investigate the risk of an infectious individual being released into the community under this regime and under alternative quarantine and testing regimes: scheduled testing of travellers only on departure from their country of origin and arrival in the destination country with no quarantine; 5-day quarantine with testing on day 3; 14-day quarantine with no scheduled testing. We also investigate the potential effect of transmission of the virus among individuals within the quarantine facility at either a moderate (*R*_*MIQ*_*=* 0.1) or high (*R*_*MIQ*_*=* 0.5) transmission level.

#### Departure and arrival testing only

Tests on departure from the home country and on arrival in the destination country with no mandatory quarantine period mean that approximately 50% of infected travellers will test positive either on departure or on arrival; the remainder will be released into the community (Figure 2A). This assumes that the time between pre-departure testing and arrival testing is 1 day. This very low detection rate reflects the high probability of a false negative test during the first five days after infection. These individuals are also likely to be highly infectious (i.e. within 5 days of exposure) when they enter the community (Figure 2B). Under this regime, there is only a very limited time window (assumed to be 1 day) for transmission while travelling. Therefore, the level of internal transmission makes little difference. This shows that a testing-only policy without quarantine would pose a very high risk to the community.

**Figure 2:**
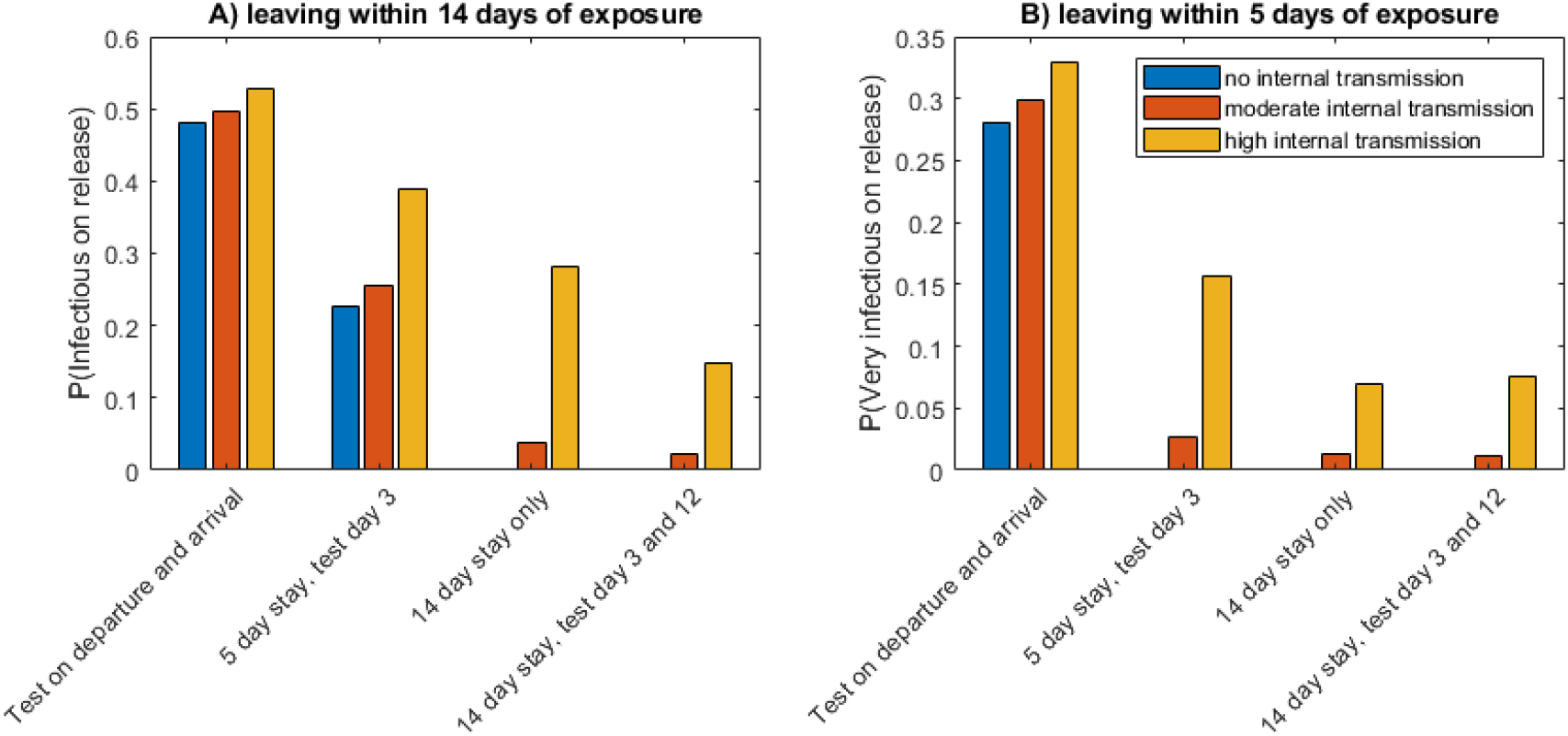
The probability per infected arrival of an individual being released into the community whilst A) infectious (within 14 days of exposure) or B) very infectious (within 5 days of exposure) under four different quarantine and testing policies, and for different rates of transmission within quarantine facilities: no transmission (*R*_*MIQ*_*=* 0), moderate transmission (*R*_*MIQ*_ *=* 0.1), and high transmission (*R*_*MIQ*_*=* 0.5).

#### 5-day quarantine with one test

A stay of 5 days in quarantine facilities with a test on day 3 reduces the probability of a highly infectious individual entering the community relative to the testing-only policy. With no transmission or moderate transmission within quarantine, around 25% of infected arrivals will leave quarantine while infectious (Figure 2A). This still poses a very high risk to the community: around one in four infected arrivals will still be infectious on leaving quarantine and up to 1 in 35 will be very infectious (Figure 2B). If there is high transmission within the quarantine facility this probability increases to around 38% of infected arrivals being infectious when leaving quarantine and over 15% being highly infectious.

#### 14-day quarantine with no scheduled testing

With a 14-day quarantine period, the probability of an infectious person leaving quarantine is 4% or 28% under moderate or high transmission respectively (Figure 2A). Under moderate transmission, this is comparable to 14 days with two tests. However, if internal transmission rates are high the additional testing gives a large decrease in risk. The risk of a very infectious person leaving is not significantly changed by removing the two tests from the 14-day quarantine period (Figure 2B).

#### 14-day quarantine with two tests

Assuming moderate transmission within quarantine facilities, the risk of an infectious individual being released into the community is around 2% (Figure 2A). The risk of a highly infectious individual (within 5 days of exposure) leaving quarantine is 1% (Figure 2B). This is the probability per infected arrival, i.e. it is expected that one highly infectious individual will be released for every 100 infected arrivals on average. Assuming high transmission due to mixing within quarantine, the risk of a highly infectious person entering the community increases to 7% per infected arrival. If there is no transmission within quarantine facilities, all individuals are no longer considered infectious (i.e. they were exposed more than 14 days ago) when leaving.

In each of the testing and quarantine regimes investigated above, the individuals who are released into the community while infectious will comprise both clinical and subclinical infections. The proportion of released infectious individuals who are subclinical will be higher than the proportion of all infections that are subclinical (assumed to be 33%). This is because clinical cases are more likely to be detected prior to being released, as they may receive an additional test triggered by symptom onset and are more likely to return a positive result when tested. Because subclinical infections are assumed to be less infectious than clinical cases (14) (16), the risk of onward transmission in the community posed by recent arrivals is less than it would be if they consisted of 33% subclinical infections. We do not attempt to model this in more detail and note that a bias towards identification of clinical cases is a common feature of screening programmes that use scheduled and/or symptom-triggered testing.

#### Measuring the risk of transmission within quarantine facilities

Under the 14-day quarantine regime with scheduled tests on day 3 and day 12, the fraction of cases detected in quarantine that are detected in the second week is a measurable indicator of potential transmission within quarantine facilities (Figure 3). When there is no within-quarantine transmission (*R*_*MIQ*_ *=* 0), around 17% of cases detected in quarantine are detected in the second week. This is the proportion of cases that return a false negative test result on day 3 and are not detected as a result of symptom onset in the first week. If this fraction increases substantially, then procedures for infection prevention in quarantine should be reviewed. Under moderate (*R*_*MIQ*_*=* 0.1) and high (*R*_*MIQ*_ *=* 0.5) within-quarantine transmission, the proportion of cases detected in the second week is 19% and 28% respectively.

**Figure 3.**
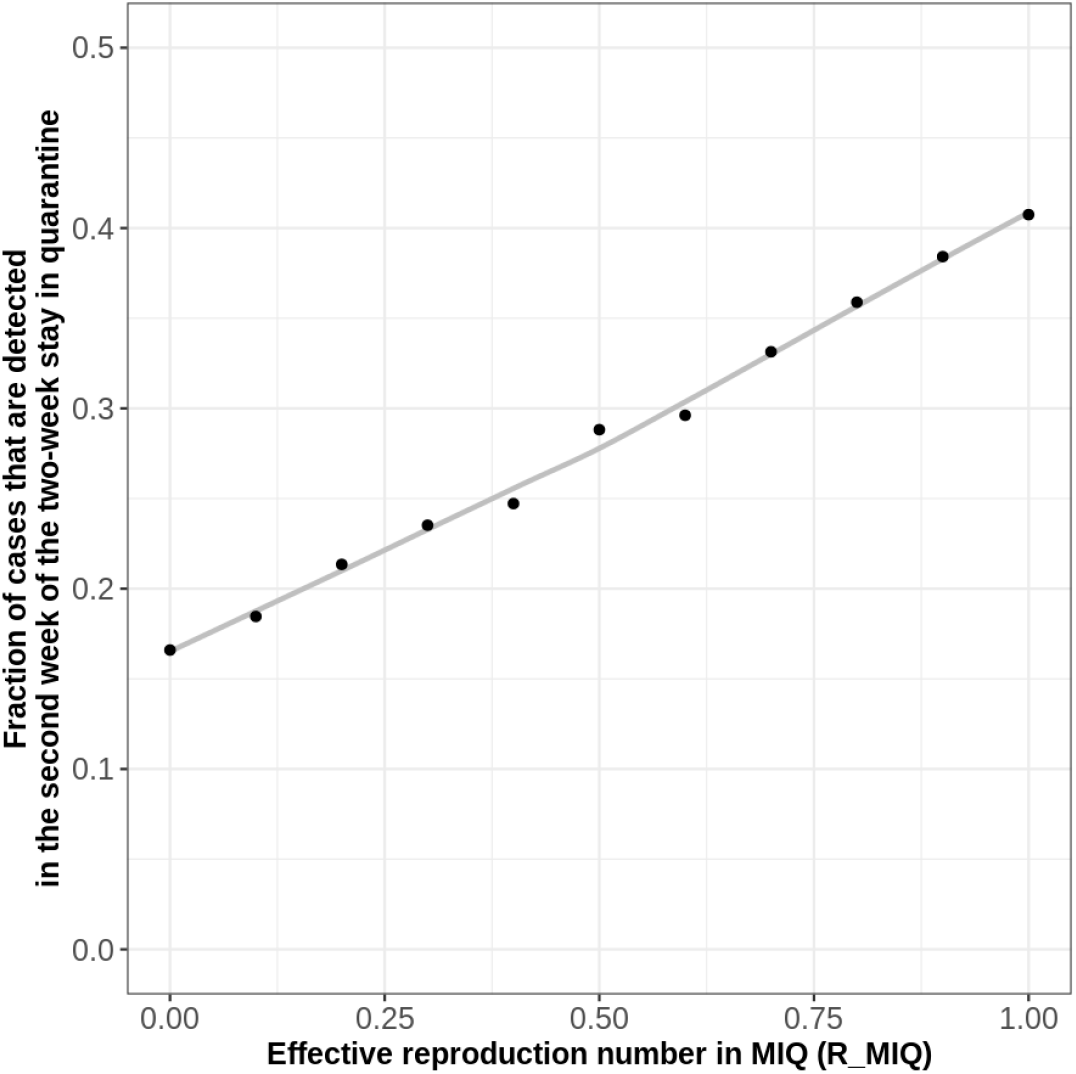
The proportion of cases detected in quarantine that are detected in the second week of a two-week stay is an increasing function of the transmission rate in quarantine facilities, measured by the within-quarantine reproduction number *R*_*M IQ*_. Results are for the 14-day quarantine regime with scheduled day 3 and day 12 testing.

### Scenario 2: community outbreak seeded by a frontline worker

We consider four testing regimes of increasing stringency: (i) no regular testing of frontline workers; (ii) fortnightly testing; (iii) weekly testing; and (iv) weekly testing with symptom checks. In all cases, frontline workers are assumed to have increased awareness of COVID-19 symptoms relative to the rest of the population. This is reflected in a shorter average time (2 days) between symptom onset and testing for those that get tested. The addition of symptom checks at the time of testing decreases the chance of a false negative for clinical cases in frontline workers more than 8 days after exposure and guarantees a positive result for any test on a clinical case in a frontline worker more than 10 days after exposure. This models diagnosis as a probable case based on clinical symptoms in the absence of a positive RT-PCR test result. For each of these four testing programs, we consider two different levels of community awareness: high, where *p*_*test*_ *=* 100% of clinical cases eventually present for testing some time after symptom onset, and low, where only *p*_*test*_ *=* 50% of clinical cases eventually present for testing. The latter might be the result of reduced public awareness or lack of community testing.

Figure 4 shows the expected size of the outbreak in the community when the first case is detected, for each of the four testing regimes with high and low community awareness. With high community awareness and weekly testing, it is likely that less than 3 individuals (in addition to the seed case) have been infected when the first case is detected (Figure 4A). If community awareness is low and there is no regular testing of frontline workers, this increases to 10 expected cases (in addition to the seed case) (Figure 4B).

**Figure 4:**
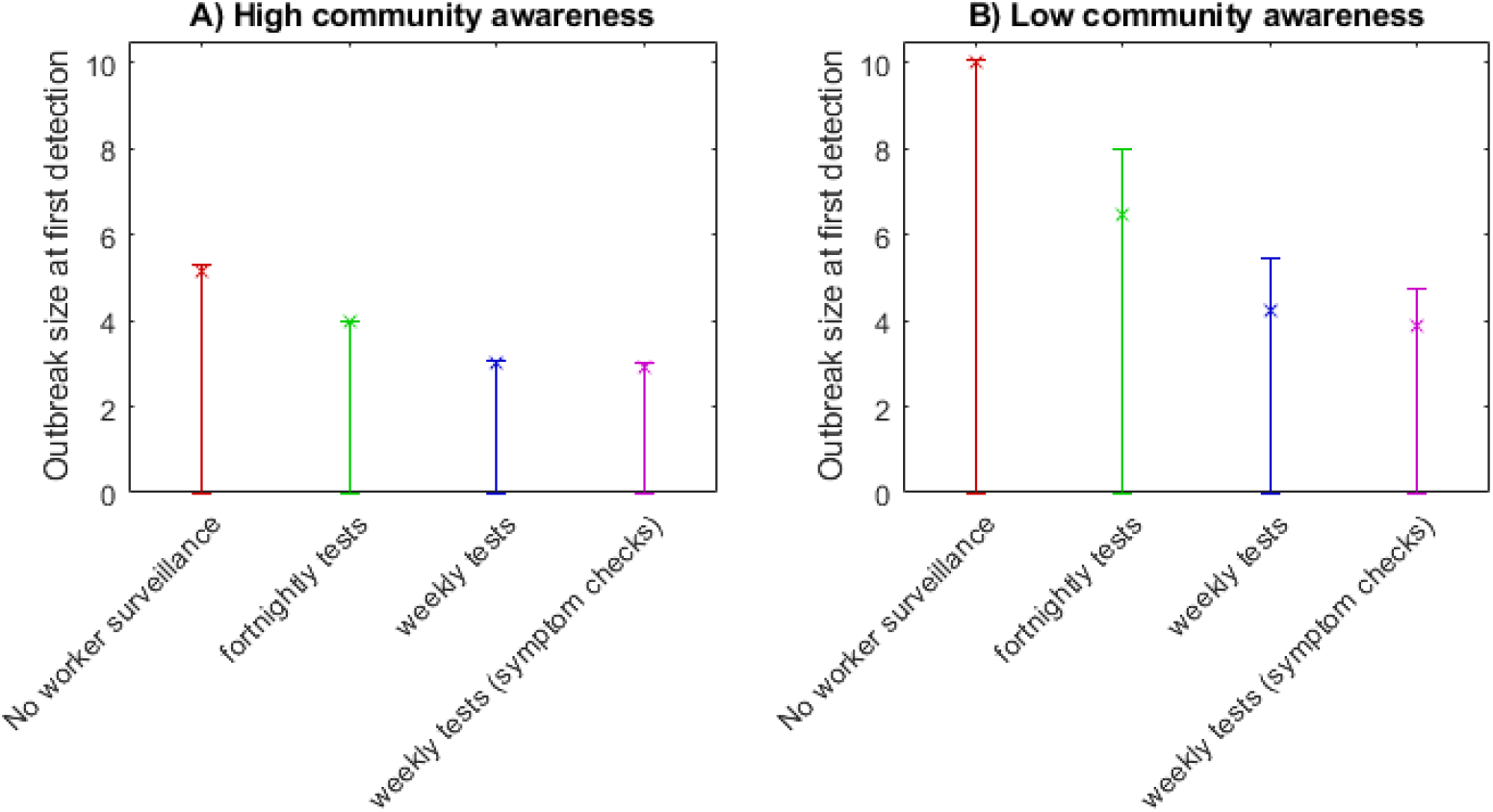
Expected number of infected individuals in the community (in addition to the seed case) when the first case is detected, under A) high community awareness and B) low community awareness. This decreases significantly with increased testing of frontline workers and improved community awareness. Expected value (x) and interquartile range from 10,000 simulations (error bars) are shown. Note that all error bars extend down to zero, meaning that at least 25% of simulations produced zero additional infections.

For each testing regime, we also record the proportion of simulations in which the outbreak: (i) dies out before being detected; (ii) is first detected in the initial frontline worker (seed case); or (ii) is first detected in a secondary case or later (Figure 5).– As the frequency of frontline testing increases, it becomes more likely that an outbreak will be detected in the seed case (>80% with the weekly testing). Decreasing community awareness, which in this scenario applies equally to frontline workers and the general population, makes little difference to the setting of the first detection, provided there is a regular frontline worker testing regime (Figure 5B). If there is no regular testing of frontline workers, the probability of the outbreak being first detected in a secondary case increases to 29% −35% under high and low community awareness, respectively.

**Figure 5:**
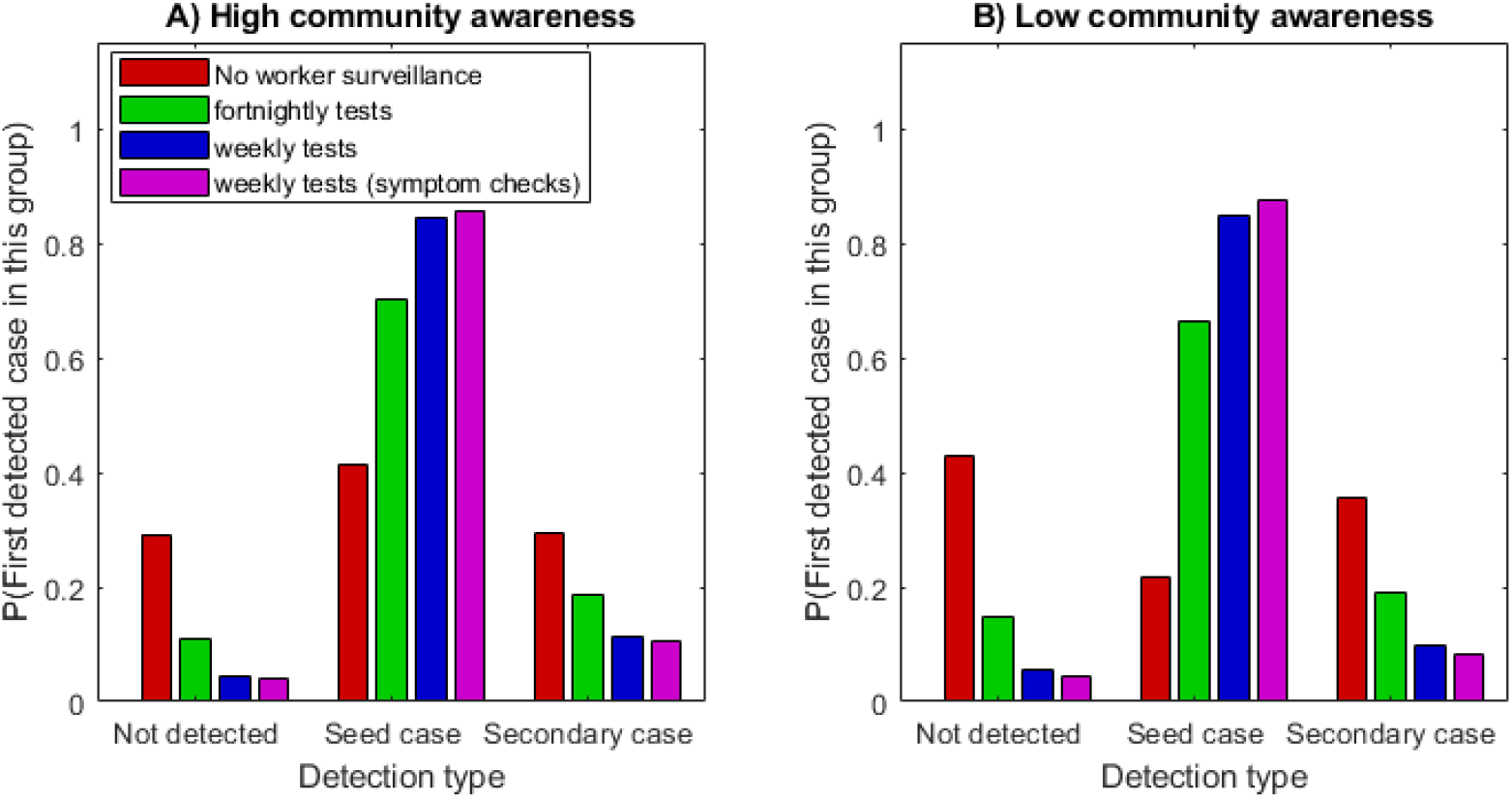
The probability of an outbreak dying out before it is detected (‘not detected’), being detected in the original frontline worker (‘seed case’) or being detected in a secondary case or later, under different testing regimes and A) high community awareness and B) low community awareness. When there is a regular testing regime for frontline workers, the probability of an outbreak being detected in the seed case is high.

The typical size of an outbreak when it is first detected depends on the time elapsed since infection of the seed case, but this information is often not immediately available. In contrast, it is usually straightforward to establish whether the first detected case is a frontline worker or not. This information can provide a valuable indicator as to the expected size of the outbreak when it is first detected. Regardless of community awareness level and testing regime, if the first detected case is a frontline worker (i.e. the seed case), the size of the outbreak is expected to be small with usually no more than 3 people infected (Figure 6). If instead the first detected case is a secondary case, including household contacts, casual contacts and those infected by environmental transmission, then the expected outbreak size is much larger. If the community awareness is high, the expected outbreak size is likely to be between 11 and 14 depending on the testing regime (Figure 6A). If community awareness is low, the expected outbreak size is likely to be between 19 and 26 (Figure 6B). The results in Figure 6 demonstrate a correlation between the type of first detected case and outbreak size. This is not a causal relationship but nevertheless provides useful information to inform the control response.

**Figure 6:**
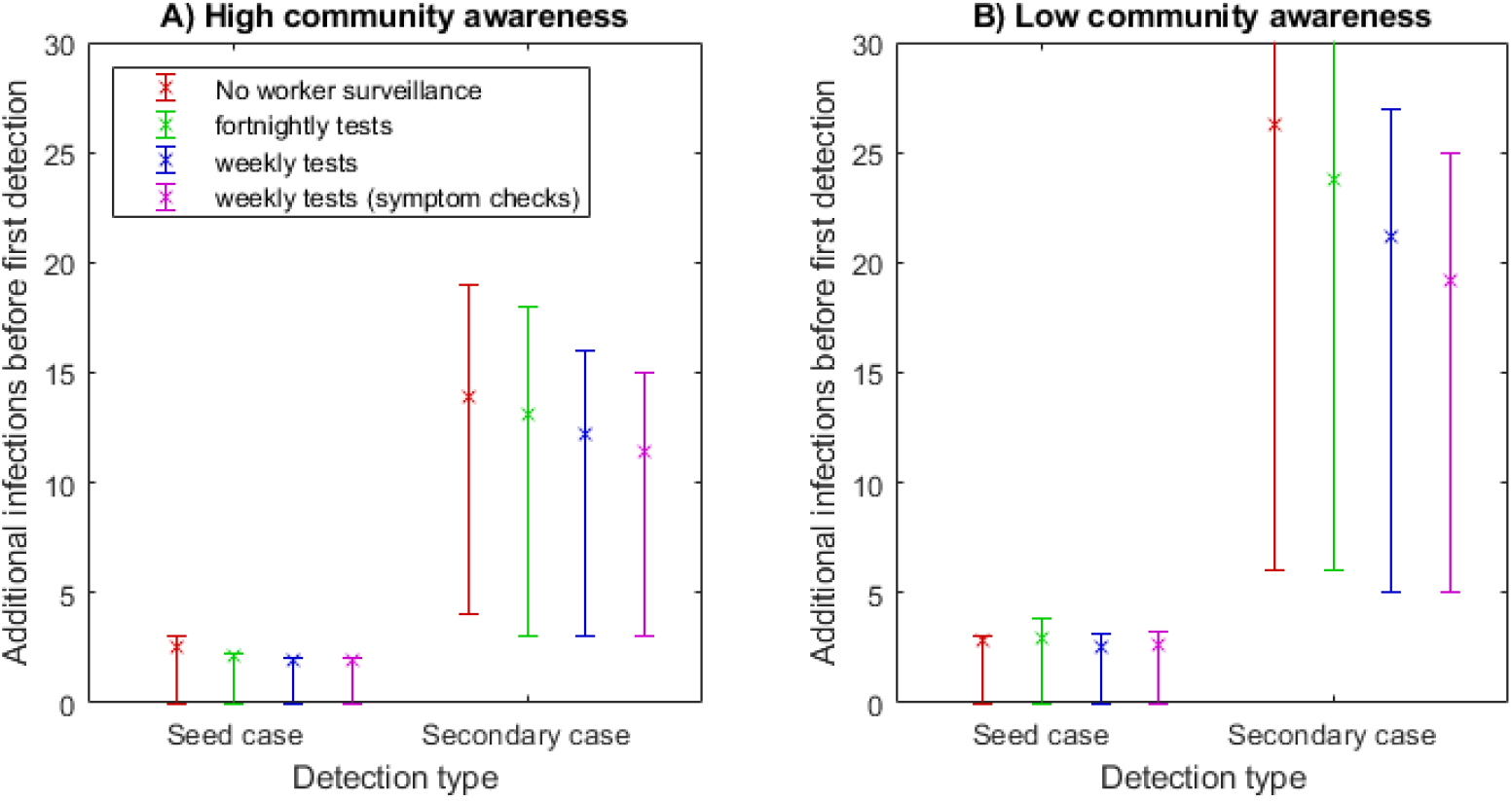
Expected number of infected individuals in the community (in addition to the seed case) when the outbreak is first detected depending on whether the it is first detected in the seed case or in a secondary or later case, under different testing regimes and A) high community awareness and B) low community awareness. Outbreaks detected in a secondary rather than seed case will be significantly larger. Expected value (x) and interquartile range from 10,000 simulations (error bars) are shown.

Small variations in the chosen parameters make very little change to the qualitative results. For example, increasing or decreasing the probability of being subclinical (25%-40%), and using a similar published infection kernel (25) gave minor numerical changes but made no difference to the overall message that weekly testing with symptom checks is a substantial improvement on no testing or even fortnightly testing. In all parameter explorations there was a substantial increase in the expected number of exposures if the first detected case was not in the frontline worker.

### Probability of inter-regional spread

We used Eq. (1) to calculate the probability that an infectious case has travelled outside a given region. These results can be used to inform decisions about travel restrictions or the geographical extent of other control measures. Figure 7 shows the risk of an infected individual having travelled outside a defined area in which the outbreak was first detected as a function of the per capita travel rate from that defined area. The risk increases with the per capita travel rate and is higher if the outbreak is not first detected in a frontline worker or if community awareness is low. To demonstrate the use of this model, Figure 7 shows illustrative travel rates for three regional scenarios for New Zealand: travel from the South Island to the North Island; travel from the North Island to the South Island; and travel from the Auckland region to the rest of New Zealand. We chose these scenarios because of the possibility of applying different control measures at a regional level and restricting travel between regions. Travel between the North and South Islands is almost exclusively by commercial airline and ferry and travel restrictions would therefore be relatively easy to enforce. Travel in and out of the Auckland region would be more difficult to restrict, but this has recently been done in response to a community outbreak detected in the Auckland region. Exact travel volume data is not available, so the travel rates shown in Figure 7 are for illustrative purpose only.

**Figure 7.**
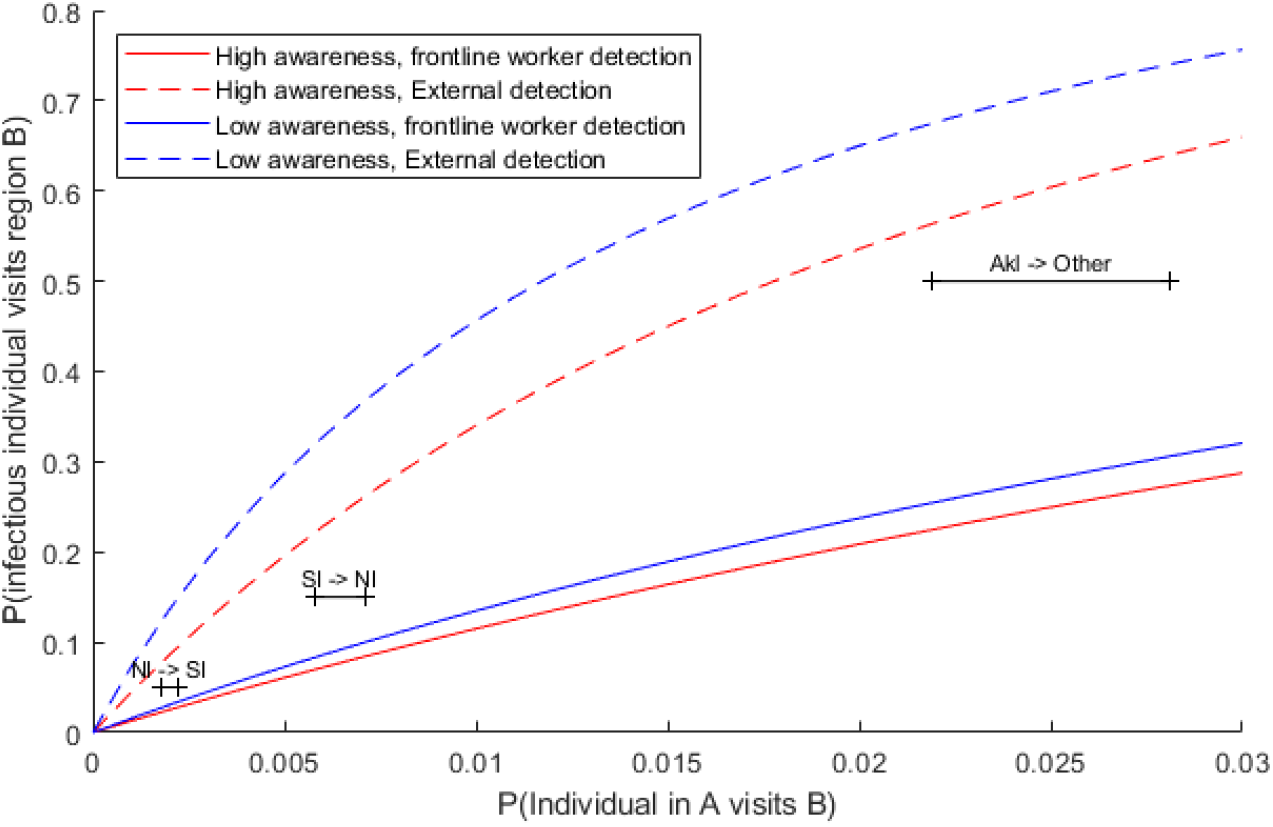
Risk of an infected individual having travelled from region A to region B before the outbreak is first detected as a function of the travel rate (daily probability of an individual in region A travelling to region B). The different curves show different levels of community awareness (red = high awareness, blue = low awareness) and different detection settings (first detected in frontline worker = solid, first detected elsewhere = dashed). Horizontal black bars indicate an illustrative range of travel rates for three New Zealand regional scenarios: outbreak detected in the North Island, risk of an infected individual having travelled to the South Island (NI -> SI); outbreak detected in the South Island, risk of an infected individual having travelled to the North Island (SI -> NI); outbreak detected in the Auckland region, risk of an infected individual having travelled outside the Auckland region (AKL -> other). Assumed travel volumes per day: between the North and South Islands 7000 – 8500; between Auckland and the rest of New Zealand 35,000 – 45,000. Population sizes: North Island 3.75 million; South Island 1.25 million; Auckland region 1.6 million.

The probability of an infected individual having travelled from the North Island to the South Island by the time an outbreak is first detected in the North Island is approximately 3% if the outbreak is first detected in a frontline worker regardless of the level of community awareness (Figure 7; solid lines within range of travel rates indicated by ‘NI -> SI’ bar). If the outbreak is not first detected in a frontline worker, the probability of travel is approximately 8% if community awareness is high, and up to 15% if community awareness is low. The probabilities of an infected individual having travelled from the South Island to the North Island by the time an outbreak is first detected in the South Island are higher than the corresponding probability of a case having travelled from the North to the South Island because of the asymmetry in population sizes. The probability of an infected individual having travelled outside the Auckland region when an outbreak is first detected there is very high, because of large travel volumes and a smaller source population (Figure 7).

## Discussion

For international arrivals at the border, the safest regime analysed was by far the 14-day stay in government-managed quarantine with two scheduled tests. Provided confirmed cases are completely isolated, and there is minimal transmission between individuals in quarantine facilities, this regime reduces the risk of an infectious case being released into the community to less than 1% per arriving case. Significantly reducing the length of stay in quarantine would increase the risk of a highly infectious individual entering the community. Relying on testing alone with no quarantine would be much higher risk, largely because of the high false negative rate of RT-PCR tests, particularly prior to symptom onset (13).

The greatest reduction in risk associated with quarantined international arrivals can be obtained by minimising mixing among guests in the facilities. This can be achieved by eliminating or carefully managing shared spaces such as smoking, exercise areas, and elevators. Evidence suggests that speaking, especially while exercising, can substantially increase the chances of transmission (26). Any contacts between guests in quarantine facilities increases the probability that someone catches the virus during their stay and is still infectious when they complete their 14-day quarantine period. Gatherings of groups of guests could allow superspreading events to occur, which would further increase the risk of infectious travellers or workers entering the community. Strict physical distancing measures within quarantine facilities are an essential component of maintaining a high probability of interception.

Our results show that the fraction of imported cases that are detected in the second week of quarantine (out of all cases detected in quarantine) is an effective indicator of the level of transmission among individuals staying in quarantine facilities. Although the exact value of this indicator is sensitive to modelling assumptions, a value of 20% or more is indicative of significant within-facility transmission. Tracking the value of this indicator over time gives a measure of the effectiveness of infection prevention in the quarantine facility, and an upward trend would point to the need to review processes.

The risk of a frontline worker inadvertently seeding a major outbreak can be significantly reduced by scheduled weekly testing combined with symptom checks by trained professionals. Clearly this should be undertaken alongside measures that minimise the risk of a frontline worker becoming infected, including training of frontline workers in infection control, supply and correct use of effective protective personal equipment at all times, cleaning potentially infected surfaces and eliminating all close contacts and shared spaces between frontline workers and quarantine guests.

The test for SARS-CoV-2 is invasive and unpleasant. Repeated compulsory testing on individuals, particularly those at relatively low risk, may be unpopular. There is also a danger of workers being reluctant to get tested or to reveal symptoms through fear of being stood down from work with no pay. This underscores the need for comprehensive paid sick leave. Our results show how valuable the efforts of those who undergo regular testing are for protecting the wider community. For instance, omitting testing of arrivals from a 14-day quarantine substantially increases the risk of an infectious individual entering the community, especially if there is a risk of significant transmission in quarantine facilities. Similarly, regular testing of frontline workers means it is far more likely that a border re-incursion is detected early, before it can grow into a large outbreak. Alternative approaches to testing that are less invasive are also being explored. For example, a recent study (27), involving over 2.6 million individuals, tested the use of a smart phone app to assess self-reported symptoms. The best fit model had a 69% positive prediction rate, implying that this method would likely miss 30% of symptomatic cases and all asymptomatic cases.. However, the symptom checker results did not state at what point in the course of the illness the model was applied, i.e. late onset symptoms may be needed to achieve this level of accuracy. So as yet, this is not likely to be a viable substitute. However, given the rapid development of antigen tests, particularly saliva-based tests, more frequent testing of frontline workers could soon be a possibility.

If a community outbreak seeded at the border is first detected outside the frontline workforce, it is likely that a large number of other infections will have occurred by the time of first detection. This applies even if the first detected case is a household contact of a frontline worker, because at this stage in the outbreak there is a high chance that transmission beyond the worker’s household has occurred. In these circumstances, the total number of cases is likely large enough that an immediate local lockdown may be necessary to bring the outbreak under control. This situation occurred in Auckland, New Zealand on 11 August 2020 and immediate local restrictions on travel, gatherings, non-essential businesses and schools were announced(28).

Other studies have also modelled the risk of transmission from imported cases and the effectiveness of border quarantine. For example, Arino, Bajeux (29) used a stochastic compartmental model to show that the rate of importations is more important than the strength of social distancing measures for determining the risk of a new localised outbreak, and that quarantine is an effective way of reducing the rate of importations. Their model predicted that the chance of 14-day quarantine successfully preventing an infected arrival entering the community while still infectious was between 70% - 90% under low and high detection rates, respectively. Hossain, Junus (30) presented a meta-population compartment model that embeds city-to-city connections and stratifies transmission into imported cases, their secondary infections and local cases. Using this model with inter-city travel data from Wuhan’s 2020 outbreak, they assessed the effectiveness of travel restrictions and quarantine for reducing the probability of an outbreak emerging in other cities. Their model assumes that quarantine prevents all onward transmission by quarantined individuals and is useful for assessing how the time taken to quarantine an infected individual affects the emergence of community outbreaks. Our model goes a step further than those of (29) and (30) by simulating transmission dynamics within quarantine facilities as well as in the community, and comparing outcomes of different quarantine and testing regimes, and different rates of within-quarantine transmission.

We used a simple model to estimate the probability that an infected individual has travelled outside the region in which the outbreak is detected. This model assumes that every individual in the region, including those infected, has an equal probability of travelling between regions each day, which ignores heterogeneity in travel patterns. Another caveat is that this method estimates the probability that an infected individual has travelled to a given region, not the probability they have passed the virus on to someone else there. The latter will depend on the time since the traveller was infected, the length of stay and contact rates in the destination region, and potential correlations among these variables. We do not attempt to quantify this, but we note the probability of onward transmission in the destination region will necessarily be less than the probability of travel, so our results provide a conservative upper bound for the risk of inter-regional spread. This method may be useful in determining the geographical scale of any response measures in response to the outbreak detection.

These findings apply to all workers who have contact with international arrivals, whether direct or indirect. Workers who experience only indirect or environmental contact with quarantined individuals have a lower overall risk of being infected, but if they are infected the consequences are the same. Maintaining elimination of COVID-19 depends on the ability to contain all cases at the border. While the controls investigated here may seem onerous, in the long run they are likely to be a preferable alternative to regional or national lockdowns.

We did not model superspreaders or superspreading events inside quarantine facilities, as these are unlikely to occur in this environment. It is possible that communal spaces and surfaces (such as buses, elevators, reception areas, door handles) could provide an avenue for environmental transmission. In our model, this would effectively correspond to an increase in the mean number of contacts parameter but is unlikely to cause superspreading events given the restrictions on individual movements. Nevertheless, communal spaces and surfaces should be regularly cleaned and good hand hygiene encouraged to minimise the possibility of environmental transmission. Supershedders (individual heterogeneity in infectiousness) can increase the risk of releasing an infectious case into the community, but this effect is small providing existing procedures are followed. We did not explicitly model families or other groups travelling together. It is possible that these groups will increase the number of cases detected in the second week because of transmission between people staying in the same room. However, for the purposes of measuring widespread transmission in quarantine this should not be considered in the fraction calculation.

## Data Availability

No new data is presented in this article

## Acknowledgements

The authors acknowledge the support of StatsNZ, ESR, and the Ministry of Health in supplying data in support of this work. The authors would also like to thank Samik Datta, Nigel French, Markus Luczak-Roesch, Anja Mizdrak and Matt Parry for informal peer review comments on an earlier version of part of this manuscript, and two anonymous referees whose suggestions helped improve and clarify this manuscript. This work was funded by the New Zealand Ministry of Business, Innovation and Employment and Te Pūnaha Matatini, Centre of Research Excellence in Complex Systems.

