## Appendix for "Managing the risk of a COVID-19 outbreak from border arrivals"

### Appendix: Operational details for government-managed quarantine in New Zealand

#### *Managed isolation and quarantine (MIQ) facilities in New Zealand*

The following is a summary of MIQ operational details extracted from MIQ (2020a) and current in December 2020.

All arrivals to New Zealand undergo initial screening at the border and, where required, undergo a health screen including a COVID-19 symptom check and temperature assessment. All arrivals are then transferred to a MIQ facility, except for those in urgent need of hospital-level medical care who are transferred directly to hospital. Transportation of arrivals (via bus, minivan or aircraft) from the port of arrival to a MIQ facility, or transfers between MIQ facilities, must comply with the following:

- All passengers (and drivers/crew) must wear a medical face mask during transportation, and until reaching their room in the MIQ facility.
- 2 metre physical distancing should be maintained on entering/exiting the vehicle and inside the vehicle if possible.
- Basic hand hygiene should be performed on entering/exiting the vehicle and immediately after handling luggage.
- When rest stops are required, these must be located away from major traffic routes. Staff must supervise rest stop areas to prevent entry by members of the public and to ensure 2 metre physical distancing is maintained. After use, rest-stop areas must be cleaned using hospital-grade detergent/disinfectant before being re-opened to the public.
- A domestic flight transporting new arrivals to a MIQ facility must not have other passengers on board. Air crew must wear face masks and gloves. The aircraft must be cleaned after use.

At time of writing, New Zealand has 32 MIQ facilities, located on its North Island in Auckland (18), Hamilton (3), Rotorua (3) and Wellington (2), and on its South Island in Christchurch (6) (MIQ, 2020b). Facilities are either lower-risk managed isolation facilities (for arrivals who are asymptomatic, have not tested positive for COVID-19, and are not a close contact of confirmed or probable COVID-19 cases) or higher-risk quarantine facilities (for confirmed or probable COVID-19 cases and their close contacts) or dual-use facilities, which have a designated and clearly delineated quarantine zone.

A close contact is defined as any person who is exposed to (within 2 metres for at least 15 minutes) or a household member of a confirmed or probable case during the case's infectious period, without appropriate personal protective equipment (PPE) (Ministry of Health, 2020). Facilities are located within hotels and satisfy the following criteria:

- Appropriate security and entry/exit points.
- Suitable room and bathroom facilities.
- Adequate provision of food and drink delivered to rooms.
- Safe laundry protocols.

- Ability to ensure people's wellbeing through the provision of online access and services.

Within 48 hours of arrival in a MIQ facility, arrivals undergo a health and wellbeing screen by a registered nurse, including COVID-19 symptom check and temperature assessment. All arrivals receive a welcome pack that includes information on COVID-19 and their stay in MIQ. Arrivals should isolate in their rooms as much as possible. They should perform regular hand hygiene and in shared spaces (e.g. reception areas, hallways) must maintain 2-metre physical distancing and wear a medical face mask. During their stay, arrivals can isolate within ‘bubbles’ (groups of people who are isolating together, e.g. family groups or other groups travelling together) in a single room or across two rooms in close proximity. People can interact with others in their bubble without use of face masks or physical distancing. If anyone within a bubble tests positive for COVID-19, all other members are considered close contacts and are moved to quarantine.

Routine testing of all arrivals (except infants aged under 6 months who are asymptomatic and are not a close contact of a confirmed or probable case) is conducted on around day 3 and day 12 of their stay in the MIQ facility. Additionally, all arrivals undergo daily health and wellbeing checks (in-person or via phone) by registered nurses or delegated health staff. Symptomatic COVID-19 cases undergo checks at least twice-daily. Anyone developing symptoms consistent with COVID-19 is immediately isolated to their room along with other members of their bubble, and tested as soon as possible. Similarly, close contacts of confirmed or probable cases are immediately isolated to their room. While awaiting test results and subject to approval, they are still offered opportunities for outdoor exercise and/or smoking under supervision to ensure PPE and 2-metre physical distancing is maintained. Anyone who receives a positive test is transferred to a quarantine facility or quarantine zone of a dual-use MIQ facility, along with their close contacts.

Arrivals have access to outdoor exercise at least once per day and access to outdoor smoking areas, under supervision by MIQ facility staff. Face masks must be worn (except when smoking), hand hygiene performed and at least 2-metre physical distancing must be maintained between all people at all times, except those who are isolating within the same bubble. Additional indoor exercise areas (including gym facilities, pools, saunas and spas) are not to be used.

Within all MIQ facilities, all surfaces and common areas (including lifts, hallways, stairwells, lobby and reception areas, and other areas) are regularly cleaned using hospital-grade detergents/disinfectant. Each facility implements a plan to ensure guests and staff can move around the facility while minimising or eliminating the potential risk of transmission to others in the facility. All facilities also have an infection prevention control training program in place to provide education to all staff, including comprehensive information and guidance on keeping themselves and their families safe.

For those who have completed their 14 days in managed isolation (or in quarantine and who have not tested positive), a medical examination and negative test result on day 12 is required for approval to exit the facility; all members of their bubble must also satisfy these criteria. Quarantined close contacts of a confirmed or probable case must re-start their 14 days of quarantine (beginning from the day after their last contact with the case while that case was deemed infectious) and a negative test on day 12 and medical examination is required to exit quarantine. For those in quarantine who have tested positive for COVID-19, no further testing is needed; instead, they must have spent at least 14 days total in a managed facility, have spent at least 10 days in a quarantine facility since testing positive or since symptom onset, have been free of symptoms for 72 hours, and receive approval from a qualified health practitioner.

*Workers in MIQ facilities in New Zealand*

All MIQ staff are screened prior to commencing work at the facility to ensure they are not highly vulnerable to severe illness, and undergo training to ensure they have appropriate understanding of general infection, prevention and control (IPC) requirements and PPE guidelines. Staff must undergo daily health checks by a qualified health practitioner at the start of each shift. Staff who start showing symptoms consistent with COVID-19 should immediately self-isolate, undergo testing and inform their manager. Daily health checks for staff are another necessary tool for early identification and management of potential cases. All staff should apply a principle of "low risk but not no risk" to protect themselves, their families and their communities. When not at work, staff are encouraged to maintain physical distancing where possible, record places they visit, wear a mask on public transport, self-monitor for COVID-19 symptoms and, if feeling unwell, self-isolate and arrange a test as soon as possible.

Workers are tested for COVID-19 once every 14 days in managed isolation facilities and once every 7 days in quarantine facilities. This includes anyone who works at a MIQ facility and those transporting people required to be in isolation or quarantine to or from a facility. Any worker who develops symptoms consistent with COVID-19 should immediately self-isolate and be tested. Workers who are a close contact of a confirmed or probable case are expected to self-isolate for 14 days. Family members or close contacts of workers who are deemed close contacts of a confirmed or probable case are notified if further testing and/or self-isolation is required. All close contacts of a worker who tests positive are self-isolated for 14 days.
